# Dosimetric advantage of ring-mounted Halcyon linac in radiation therapy for lung tumors

**DOI:** 10.1101/2025.08.19.25334035

**Authors:** Shuangyan Yang, Yaping Xu

**Affiliations:** Department of Radiotherapy, Shanghai Pulmonary Hospital, Tongji University, Shanghai, 200433, P.R. China

**Author notes:** Corresponding Author: Dr. Yaping Xu, Department of Radiotherapy, Shanghai Pulmonary Hospital, Tongji University, Shanghai, 200433, P.R. China. **Fund programs:** Shanghai Science and Technology Program (21DZ2201900); Bethune Young and Middle-aged Physician Oncology Research Fund (YDTR-008); Shanghai Pulmonary Hospital National Natural Cultivation Project(fkzr2516).

**Keywords:** Lung cancer, Intensity-modulated radiotherapy, Dosimetric comparation, Halcyon, TrueBeam

## Abstract

**Objective:** To systematically compare the dosimetric performance of two radiotherapy devices, Halcyon and TrueBeam, in intensity-modulated radiotherapy (IMRT) plans for non-small cell lung cancer (NSCLC), providing a reference for hospitals to optimize NSCLC radiotherapy based on different devices.

**Materials and Methods:** A retrospective analysis was conducted on 102 NSCLC patients who received curative radiotherapy. All initial plans were designed on the Eclipse 15.6 system based on TrueBeam parameters (6MV FF photon beam, IMRT, dose grid of 2.5mm). New plans were re-optimized using Halcyon parameters while maintaining the same field directions and other parameters. The comparison indicators included: planning target volume (PTV) dose metrics (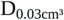, D_2%_, D_98%_, D_mean_, V_98%_, conformity index CI, homogeneity index HI); dose metrics for organs at risk (OARs) (Lungs: V_5_-V_60_, MLD; Heart: V_5_-V_60_, D_mean_; remaining normal tissues NT: V_5_-V_30_); monitor units (MUs) counts and plan complexity index (PCI). Differences were analyzed using paired t-tests or Wilcoxon tests in SPSS 27.0, and the correlation between MUs and PCI was assessed.

**Results:** The PTV conformity index (CI) of Halcyon plans was significantly superior to that of TrueBeam (0.75±0.12 vs. 0.73±0.12, P<0.001). Halcyon’s 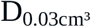 was higher than TrueBeam (6555.20±117.50 cGy vs. 6525.20±99.38 cGy, P<0.001), and V_98%_ was slightly lower than TrueBeam (95.83±11.74% vs. 95.99±11.38%, P=0.046). There were no statistically significant differences in D_2%_, D_98%_, D_mean_, and HI (P>0.05). Halcyon significantly reduced the volume of lungs, heart, and other normal tissues exposed to low and intermediate doses and lowered the mean lung dose (P<0.001). Halcyon’s MUs were significantly higher than TrueBeam’s (1323.1±374.91 vs. 1121.1±384.84, P<0.001), but its PCI was significantly lower than TrueBeam’s (0.09±0.03 vs. 0.17±0.05, P<0.001), indicating that Halcyon plans are more complex. The correlation between PCI and MUs was weak for Halcyon (R^2^=0.07), while the correlation was stronger for TrueBeam (R^2^=0.54).

**Conclusion:** Compared to TrueBeam, Halcyon demonstrated significant dosimetric advantages: superior target volume dose conformity and effective reduction in the volume of normal tissues (lung, heart, and remaining tissues) exposed to low and intermediate doses, especially in large PTV volumes where low-dose protection for NT was more significant. Although Halcyon plans are more complex and involve higher MUs, the complexity has a lower dependency on MUs, showing greater stability. Halcyon offers a safer treatment device choice for lung cancer patients who require precise dose control and low-dose protection.

## Introduction

Lung cancer remains the leading cause of cancer-related deaths worldwide, with approximately 2.48 million new cases and 1.8 million deaths reported in 2022[1]. Radiotherapy plays a key role in the treatment of lung cancer, particularly for patients who are inoperable or require combined therapy. The advent and widespread application of advanced techniques such as intensity-modulated radiotherapy (IMRT) and stereotactic body radiotherapy (SBRT) have significantly improved the ability to escalate tumor dose while minimizing radiation to adjacent normal tissues[2]. However, thoracic radiotherapy faces numerous challenges, including the the complex impact of respiratory motion on treatment planning and delivery[3], as well as the potential for radiation-induced damage to normal lung tissue.

Radiation-induced lung injury (RILI), encompassing conditions such as radiation pneumonitis and pulmonary fibrosis, remains a major dose-limiting factor in lung cancer radiotherapy. It affects approximately 5% to 20% of patients, severely impacting their quality of life and overall prognosis[4]. Additionally, radiation-related cardiac toxicity has emerged as an important concern, with evidence indicating a close association between increased heart dose and decreased survival rates[5]. Consequently, current research in lung cancer radiotherapy focuses not only on enhancing target dose conformity and uniformity but also on reducing low-to-intermediate dose exposure to normal organs, particularly the lungs and heart[6][7].

The choice of radiotherapy equipment is a key factor influencing final dosimetric outcomes, and the clinical performance of different linear accelerators (Linacs) requires thorough evaluation. Halcyon and TrueBeam are two widely used platforms from Varian Medical Systems, but they have substantial design differences. Halcyon adopts an innovative O-ring gantry design equipped with a dual-layer, staggered, stacked multileaf collimator (MLC): two layers of 1 cm-wide leaves offset by 0.5 cm, forming an MLC system with an equivalent modulation resolution of 5 mm[8]. This system omits the use of jaws and keeps interleaf transmission below 0.4% of the primary beam—significantly lower than conventional MLCs (e.g., TrueBeam’s Millennium MLC or HD-MLC, with leakage around 1.5%)[9]. This unique MLC design, combined with the high-speed gantry rotation capability, offers potential dosimetric advantages for Halcyon, especially in reducing doses to organs at risk (OARs). Studies have shown that, compared with TrueBeam, Halcyon plans can significantly reduce certain OAR doses in various clinical scenarios (e.g., spinal irradiation, breast cancer, lung SBRT), such as lowering spinal cord V_10Gy_ and V_18Gy_ values, reducing contralateral breast maximum dose, lowering maximum and specific-volume doses to the ribs and heart (e.g., rib Dmax, D1cc; heart Dmax), and potentially reducing low-dose lung irradiation volumes (e.g., V_5Gy_)[10][11][12]. Furthermore, Halcyon’s highly automated “one-step” workflow, although possibly associated with increased MUs, can significantly shorten overall treatment time, potentially reducing intrafraction motion errors and improving patient comfort[12].

Although both Halcyon and TrueBeam have been widely adopted in clinical practice, large-scale, systematic dosimetric comparisons for lung cancer remain relatively limited. Given that precise dose delivery and effective normal tissue protection are critical to the efficacy and safety of lung cancer RT, a comprehensive evaluation of these two platforms in lung cancer treatment is essential for optimizing equipment selection and treatment planning strategies. This study retrospectively analyzed IMRT planning data from 102 patients with non-small cell lung cancer (NSCLC) to systematically compare the dosimetric differences between Halcyon and TrueBeam in terms of planning target volume (PTV) coverage, OAR sparing (including lungs, heart, and other critical structures), total MU, and plan complexity index (PCI). In addition, this study explored the correlations between dosimetric parameters and clinical variables (such as tumor location and volume), with the aim of identifying key factors influencing plan quality and providing evidence-based guidance for clinical decision-making and equipment utilization.

## Materials and methods

### Patient selection and treatment planning

This retrospective study included 102 patients with NSCLC who underwent definitive radiotherapy at the Department of Radiotherapy, Shanghai Pulmonary Hospital, affiliated with Tongji University, between January 2021 and July 2024. The median age of the patients was 66 years (range: 24–91), with 88 males (86.3%) and 14 females (13.7%). Distribution of T staging: 44 patients (43.1%) with T1–2 stage and 56 patients (54.9%) with T3–4 stage. Distribution of N staging: 23 patients (22.5%) with N0–1 stage and 79 patients (77.5%) were N2–3. The median gross tumor volume (GTV) was 35.89 cm^3^ (range: 2.61–426.27 cm^3^), and the median planning target volume (PTV) was 169.65 cm^3^ (range: 68.30–697.37 cm^3^). Distribution of PTV volume: 61 patients (59.8%) in the 0–200 cm^3^ group, 33 patients (32.4%) in the 201– 400 cm^3^ group, and 8 patients (7.8%) in the >400 cm^3^ group. All initial radiotherapy plans were made using TrueBeam machine parameters on the Eclipse 15.6 treatment planning system (Varian Medical Systems, USA). The plans were optimized using 6 MV flattening filter (FF) photon beams for fixed-field IMRT, with a dose calculation grid size of 2.5 mm × 2.5 mm × 2.5 mm. The prescribed dose was 60 Gy in 30 fractions. Dosimetric requirements included covering at least 95% of the PTV volume and 99% of the GTV volume with the prescribed dose. Dose constraints for OARs were strictly followed according to RTOG clinical trial protocols and NCCN guidelines. After maintaining the same field direction and other planning parameters, new plans were optimized using Halcyon machine parameters.

This study was conducted in accordance with the Declaration of Helsinki and its subsequent amendments, and was approved by the Ethics Board of the Shanghai Pulmonary Hospital (No. K25-572). The requirement of individual consent for this retrospective analysis was waived. Data for this retrospective analysis were collected and analyzed between May 2025 and July 2025. The patient data was completely anonymous, and during or after data collection, the authors could not obtain information that could identify individual participants.

### Evaluation metrics

The evaluation metrics for PTV included: dose to the 0.03 cm^3^ volume of PTV (D_0.03cc_), dose to the 2% volume (D_2%_), dose to the 98% volume (D_98%_), mean dose (D_mean_), the percentage of volume receiving 98% of the prescribed dose (V_98%_), conformity index (CI), and homogeneity index (HI). To assess the differences in radiation exposure for OARs under different doses, the main OARs evaluated were the lungs and heart. The dose metrics included: the percentage of volume receiving doses of 5, 10, 15, 20, 25, 30, 40, 50, and 60 Gy (V_5_, V_10_, V_15_, V_20_, V_25_, V_30_, V_40_, V_50_, V_60_) and the mean dose. Additionally, the remaining normal tissue outside the PTV, defined as the outer contour minus a 2.5 cm expansion of the PTV, was also included in the evaluation, with dose metrics including: the percentage of volume receiving doses of 5, 10, 15, 20, 25, and 30 Gy (V_5_, V_10_, V_15_, V_20_, V_25_, V_30_). Moreover, PCI was analyzed in relation to MUs for both devices. The CI and HI of PTV were calculated using the following formulas[13]:

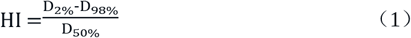

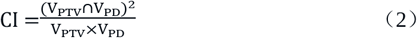

where V_PTV_ denotes the PTV volume, and V_PD_ is the volume covered by the prescription dose (PD). The PCI is a comprehensive evaluation metric based on the MUs and relevant geometric parameters of the fields, reflecting the complexity of beam modulation. The formula for PCI is as follows[14]:

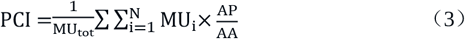

where i represents the i-th control point, N is the total number of control points, MU_i_ is the machine monitor for the i-th control point, AP is the aperture perimeter, and AA is the aperture area. A smaller PCI value indicates a more complex plan. PCI values were calculated using Python on the exported DICOM files. This study focused on the correlation between PCI and MUs, and thus did not delve into the specific methods of obtaining other parameters.

### Statistical analysis

Statistical analysis was performed using SPSS 27.0 software. Continuous variables were expressed as mean ± standard deviation (mean ± SD). The normality of data was assessed using the Shapiro-Wilk test. For normally distributed data, paired t-tests were used to compare dosimetric differences; for non-normally distributed data, the Wilcoxon signed-rank test was applied. Pearson and Spearman correlation coefficients were used to analyze the correlation between the dose deviation of OARs and PTV volume for the two radiotherapy devices. Linear regression was performed for MUs and PCI, with the regression coefficient R^2^ indicating the strength of the correlation; a larger R^2^ indicates a stronger linear relationship. All statistical tests were two-sided, and a P-value of <0.05 was considered statistically significant.

## Results

### Dosimetric Deviations of PTV

Table 1 summarizes the dosimetric parameters for the target volumes of 102 NSCLC patients treated with Halcyon and TrueBeam radiotherapy devices. In terms of PTV dose metrics, Halcyon showed a higher maximum dose (D_0.03cc_) than TrueBeam, and the difference was statistically significant (t = 3.53, P < 0.001). Regarding dose coverage, the V_98%_ of Halcyon was slightly lower than that of TrueBeam (t = -2.29, P = 0.046). For other dose parameters (such as D_2%_, D_98%_, and D_mean_), the differences between the two groups were small, and no significant changes were observed (P > 0.05). In terms of dose conformity, Halcyon plans demonstrated a significant advantage (t = 5.41, P < 0.001). However, no statistically significant differences were found between the two groups in terms of the homogeneity index (HI) (P > 0.05).

**Table 1.** Comparison of PTV dosimetric parameters between Halcyon and TrueBeam plans.

| Metrics | Halcyon | TrueBeam | t | P |
| --- | --- | --- | --- | --- |
| $D_{0.03cc}$(cGy) | 6555.20±117.50 | 6525.20±99.38 | 3.53 | $<0.001$ |
| $D_{2\%}$ (cGy) | 6439.30±109.93 | 6434.90±91.86 | 0.48 | 0.63 |
| $D_{98\%}$ (cGy) | 5853.30±144.90 | 5856.40±157.03 | -0.75 | 0.31 |
| $D_{mean}$ (cGy) | 6218.30±110.12 | 6215.80±105.91 | 0.42 | 0.44 |
| V<sub>98%</sub> (%) | 95.83±11.74 | 95.99±11.38 | -2.29 | 0.046 |
| HI | 0.09±0.02 | 0.09±0.03 | 0.58 | 0.37 |
| CI | 0.75±0.12 | 0.73±0.12 | 5.41 | <0.001 |

### Dosimetric Deviations of OARs

Table 2 presents the dosimetric metrics for OARs under Halcyon and TrueBeam radiotherapy devices. In terms of lung dose sparing, Halcyon plans demonstrated significantly lower values across most dose volume parameters (P < 0.001), particularly for key metrics such as V_5_, V_10_, V_15_, and V_20_, where the t-values were negative with large absolute values, indicating that Halcyon was more effective in reducing low-to-medium radiation dose exposure to normal lung tissue. Additionally, the MLD for Halcyon was significantly lower than that of TrueBeam (t = -6.91, P < 0.001), further confirming its advantage in reducing lung radiation exposure.

**Table 2.** Comparison of OARs dosimetric parameters between Halcyon and TrueBeam plans.

| Metrics | Halcyon | TrueBeam | t | P |
| --- | --- | --- | --- | --- |
| Lung |  |  |  |  |
| V<sub>5</sub> (%) | 29.42±8.07 | 31.04±8.621 | -10.87 | <0.001 |
| V<sub>10</sub> (%) | 22.95±6.13 | 24.01±6.57 | -10.05 | <0.001 |
| V<sub>15</sub> (%) | 19.53±5.08 | 20.30±5.33 | -8.74 | <0.001 |
| V<sub>20</sub> (%) | 16.95±4.38 | 17.56±4.55 | -7.74 | <0.001 |
| V<sub>25</sub> (%) | 14.67±3.85 | 15.05±3.95 | -5.67 | <0.001 |
| V<sub>30</sub> (%) | 12.30±3.44 | 12.65±3.53 | -5.54 | <0.001 |
| V<sub>40</sub> (%) | 7.51±2.78 | 8.01±2.88 | -8.29 | <0.001 |
| V<sub>50</sub> (%) | 4.16±1.99 | 4.44±2.12 | -8.54 | <0.001 |
| V<sub>60</sub> (%) | 1.68±1.10 | 1.71±1.08 | -1.81 | 0.009 |
| MLD (cGy) | 904.76±227.60 | 929.16±237.39 | -6.91 | <0.001 |
| Heart |  |  |  |  |
| V<sub>5</sub> (%) | 25.34±22.36 | 27.85±23.55 | -8.28 | <0.001 |
| V<sub>10</sub> (%) | 20.16±19.18 | 22.25±20.47 | -7.24 | <0.001 |
| V<sub>15</sub> (%) | 16.71±15.99 | 18.38±17.03 | -6.03 | <0.001 |
| V<sub>20</sub> (%) | 14.19±13.73 | 15.60±14.63 | -5.29 | <0.001 |
| V<sub>25</sub> (%) | 12.08±11.84 | 13.23±12.46 | -4.21 | <0.001 |
| V<sub>30</sub> (%) | 10.04±10.30 | 10.92±10.57 | -3.54 | <0.001 |
| V<sub>40</sub> (%) | 5.79±6.83 | 6.40±6.77 | -3.10 | <0.001 |
| V<sub>50</sub> (%) | 2.82±3.74 | 3.08±3.65 | -2.98 | <0.001 |
| V<sub>60</sub> (%) | 1.02±1.49 | 1.07±1.52 | -1.78 | <0.001 |
| D<sub>mean</sub> (cGy) | 769.31±629.83 | 819.25±661.40 | -4.73 | <0.001 |
| NT |  |  |  |  |
| V<sub>5</sub> (%) | 11.00±3.84 | 11.69±4.17 | -11.23 | <0.001 |
| V<sub>10</sub> (%) | 8.23±2.85 | 8.73±3.11 | -11.49 | <0.001 |
| V<sub>15</sub> (%) | 6.46±2.30 | 6.83±2.47 | -9.79 | <0.001 |
| V<sub>20</sub> (%) | 5.16±1.91 | 5.41±2.03 | -8.41 | <0.001 |
| V<sub>25</sub> (%) | 3.99±1.56 | 4.14±1.66 | -5.55 | <0.001 |
| V<sub>30</sub> (%) | 2.89±1.27 | 2.95±1.34 | -2.39 | <0.017 |

In terms of heart dose protection, Halcyon plans also exhibited lower dose volume values, with all differences being statistically significant (P < 0.001). Particularly in the low-to-medium dose range of 5–20 Gy, the t-values were negative with large absolute values, indicating that Halcyon had an advantage in reducing low-to-medium radiation exposure to the heart. Further analysis revealed that the mean heart dose for Halcyon was lower (t = -4.73, P < 0.001), which helps in reducing heart radiation exposure.

For the protection of remaining normal tissue, Halcyon plans showed lower values than TrueBeam across several key dose volume parameters (such as V_5_, V_10_, V_15_, V_20_, V_25_, and V_30_), and all differences were statistically significant (P < 0.05). Particularly in low-to-medium dose volume parameters like V_5_, V_10_, V_15_, and V_20_, Halcyon demonstrated better dose control, with negative t-values and large absolute values, indicating that Halcyon effectively reduces radiation exposure to normal tissues.

Figure 1 illustrates the mean dose-volume histograms (DVHs) for the lungs, heart, and remaining normal tissue (NT) under the Halcyon and TrueBeam systems. Halcyon showed lower volume percentages in the low-dose regions, with narrower IQR ranges, indicating more uniform and effective dose protection.

**Figure 1.**
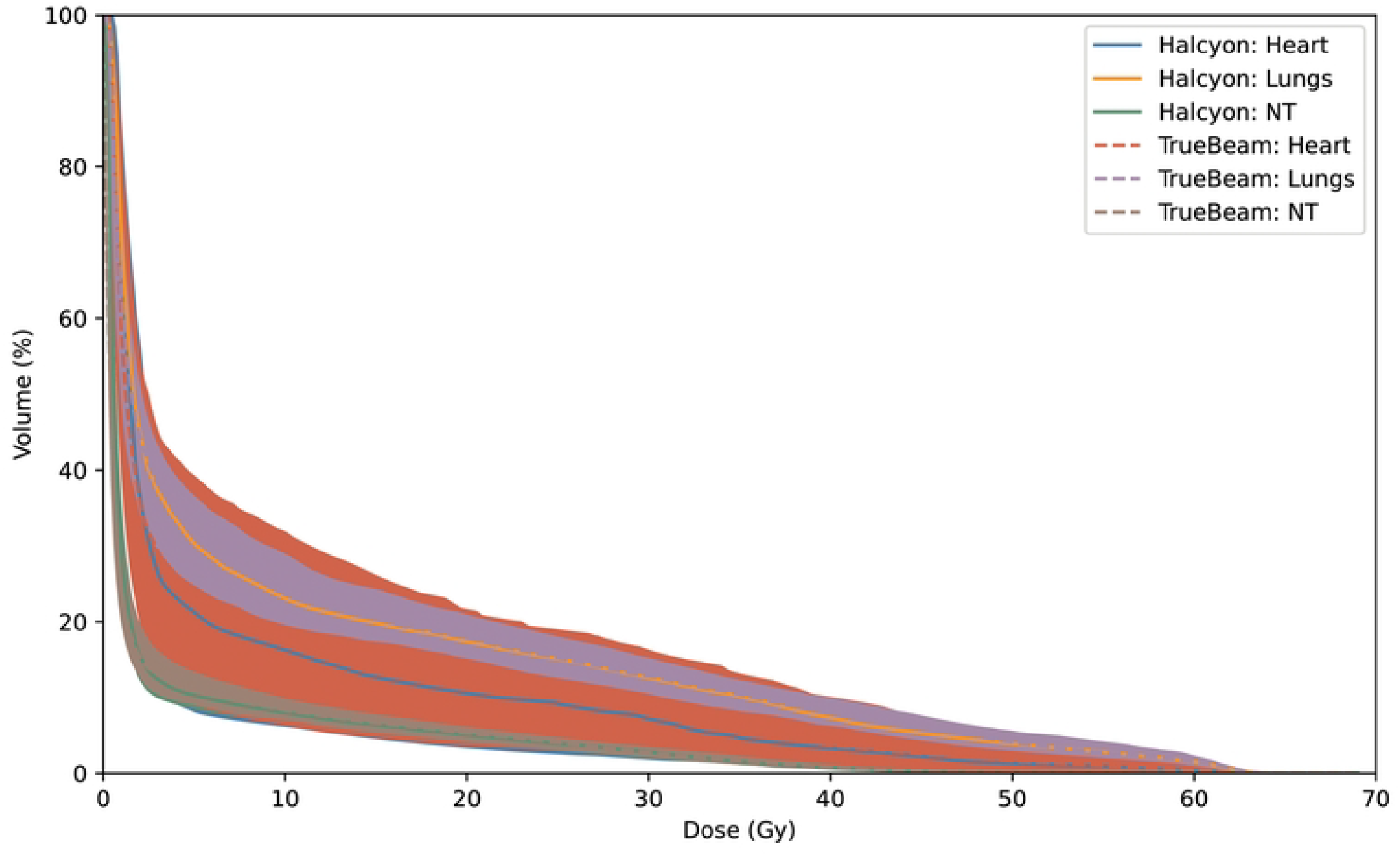
Mean DVH curves for the Heart, Lungs, and NT in Halcyon and TrueBeam plans

### Correlation Analysis Between OAR Dose Metric Deviations and PTV Volume

A further analysis was conducted to examine the correlation between deviations in OAR dose metrics and PTV volume under the Halcyon and TrueBeam systems. The correlation analysis results are shown in Table 3. Pearson and Spearman correlation analyses indicated that lung metrics such as V_5_, V_10_, and V_20_ showed no significant correlation with PTV volume (P > 0.05), suggesting that the dose distribution in the low-to-medium dose regions of the lungs remained relatively stable despite changes in PTV volume.

**Table 3.**
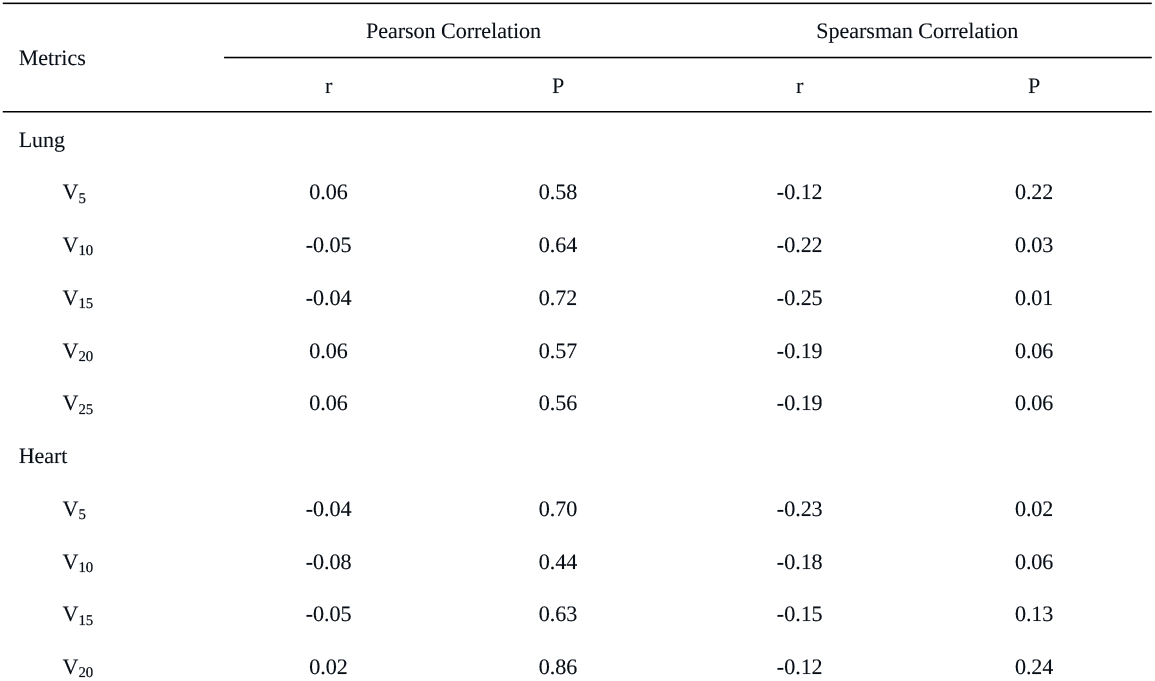
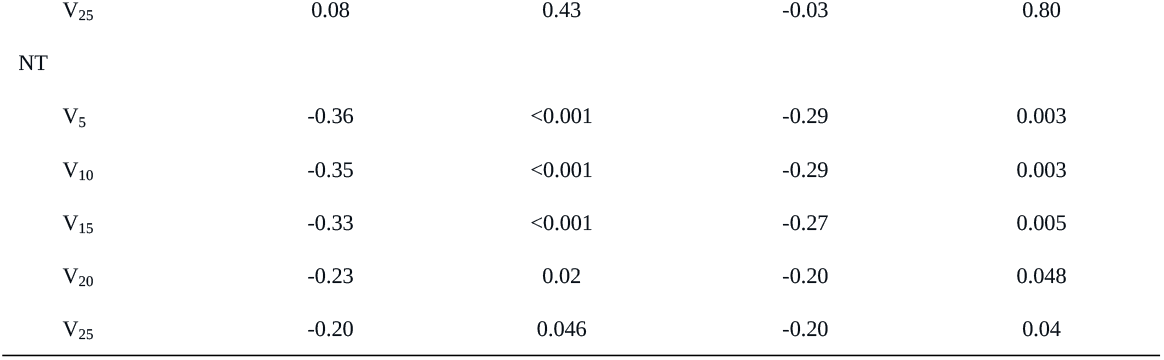
Correlation between dose metric deviation of OARs and PTV volume under the Halcyon and TrueBeam systems.

However, the Spearman correlation between the heart’s V_5_ and PTV volume was significant (P = 0.02), indicating that the heart’s low-dose region is more sensitive to changes in PTV volume. For the remaining normal tissue (NT), V_5_, V_10_, V_15_, V_20_, and V_25_ showed significant negative correlations with PTV volume (P < 0.05), with Pearson correlation coefficients ranging from -0.36 to -0.20 and Spearman correlation coefficients ranging from -0.29 to -0.20. This suggests that as PTV volume increases, the low-to-medium dose regions in NT are significantly reduced, further confirming that for larger PTVs, Halcyon provides superior low-dose control compared with TrueBeam.

### Correlation Analysis Between MUs and PCI

In the comparison of MUs and PCI, there were significant differences between Halcyon and TrueBeam. As shown in Table 4, Halcyon had significantly higher MUs than TrueBeam (t = 11.15, P < 0.001), while Halcyon had a significantly lower PCI than TrueBeam (t = -15.82, P < 0.001). Further analysis of the Pearson correlation between PCI and MUs in both systems showed that Halcyon had a weak positive correlation between PCI and MUs (r = 0.26, P = 0.009), indicating that the plan complexity in Halcyon is less dependent on MUs. On the other hand, TrueBeam exhibited a stronger positive correlation between PCI and MUs (r = 0.73, P < 0.001), suggesting that the variation in MUs in the TrueBeam system has a stronger explanatory power for plan complexity. Figure 2 shows the linear fit results of MUs and PCI under both Halcyon and TrueBeam systems.

**Table 4.** Comparison of MUs and PCI between Halcyon and TrueBeam plans.

| Metrics | Halcyon | TrueBeam | t | P |
| --- | --- | --- | --- | --- |
| MUs | 1323.1±374.91 | 1121.1±384.84 | 11.15 | <0.001 |
| PCI | 0.09±0.03 | 0.17±0.05 | -15.82 | <0.001 |

**Figure 2.**
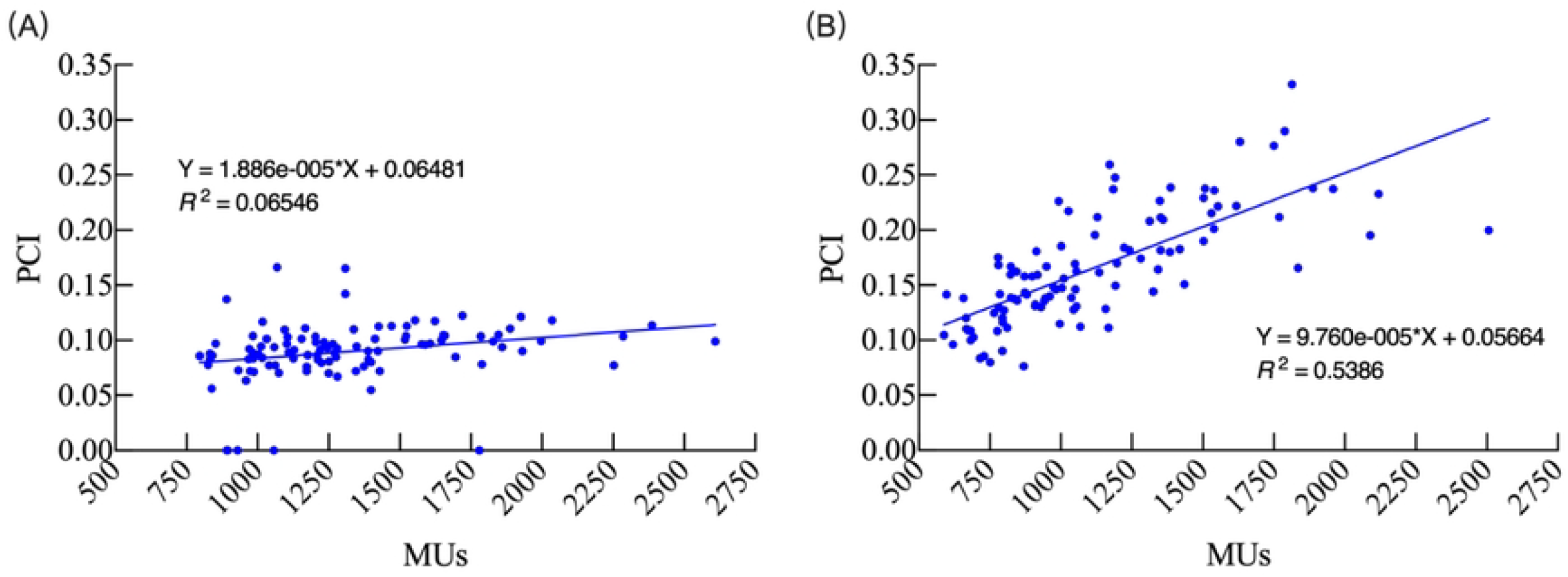
Linear fitting of MUs and PCI under the Halcyon (A) and TrueBeam (B) systems

## Discussion

This study aims to compare the dosimetric performance of two radiotherapy devices, Halcyon and TrueBeam, in radiation therapy plans for 102 NSCLC lung cancer patients, with a focus on dose conformity, OAR sparing, and treatment plan complexity. The results indicate that Halcyon demonstrated advantages in several key dosimetric indicators, particularly in dose conformity and low-dose protection.

Regarding PTV dose distribution, the CI of Halcyon was significantly higher than that of TrueBeam (P < 0.001), indicating that Halcyon performed better in matching the prescribed dose to the target shape and in controlling high-dose spill. This is consistent with literature reports that Halcyon offers superior target dose coverage and conformity, and that improved conformity helps reduce the risk of radiation exposure to surrounding normal tissues[15], as evidenced in this study by lower V_60Gy_ values for the lungs and heart in the Halcyon plans. In addition, the slightly lower V_98%_ observed with Halcyon (95.83 % vs. 95.99 %, P=0.046) is unlikely to be clinically meaningful because coverage exceeded 95% in both cohorts.

In terms of OAR protection, Halcyon showed significant advantages in the mid- and low-dose regions for the lungs, heart, and NT. Studies have indicated that lung V20 and MLD are closely related to the incidence of radiation-induced lung injury (RILI)[16], and the V10 radiation exposure volume is also considered a predictor for lung complications[17]. Compared to TrueBeam, Halcyon exhibited significantly lower lung V_5_, V_10_, V_20_, and MLD (P < 0.001), suggesting its potential advantage in reducing RILI, which has also been confirmed in studies by Pokhrel et al.[12]. This advantage may be attributed to Halcyon’s dual-layer MLC structure and flattening-filter-free (FFF) beam characteristics, which result in smaller radiation exposure volumes in the low-to-medium dose regions[12][18]. Additionally, correlation analysis revealed that Halcyon’s performance in low-dose lung protection was not significantly affected by changes in PTV volume (P > 0.05), whereas the low-to-medium dose radiation exposure of NT significantly decreased as the PTV volume increased (P < 0.05), suggesting that Halcyon can more effectively reduce low-to-medium dose radiation exposure volumes in the treatment plans for locally advanced lung cancer patients.

For heart dose protection, Halcyon outperformed TrueBeam in the volume percentages of mid- and low-dose regions (V_5_, V_10_, and V_20_) and the mean heart dose (Dmean) (P < 0.001). Studies have shown that V_5_, V_30_, and V_50_ of the heart are closely related to the incidence of cardiovascular events, and higher Dmean values are associated with decreased overall survival[19][20]. Compared to TrueBeam, Halcyon demonstrated significant advantages in V_5_, V_10_, and Dmean for the heart (P < 0.001), further proving its effectiveness in heart dose protection. Additionally, further correlation analysis showed a significant positive correlation between Halcyon’s heart V_5_ and PTV volume (P = 0.02), suggesting that the low-dose heart exposure volume might increase with larger target volumes, a phenomenon that requires further investigation in future studies.

Regarding plan complexity and machine monitor units (MUs), Halcyon’s MUs were significantly higher than TrueBeam’s (P < 0.001), but its Plan Complexity Index (PCI) was significantly lower (P < 0.001). Generally, a smaller PCI value indicates higher plan complexity, and Halcyon’s dual-layer MLC structure may increase the plan’s complexity[21]. Further analysis showed that Halcyon’s PCI had a weak linear correlation with MUs (R^2^ = 0.07), suggesting that its plan complexity is less dependent on machine monitor units, likely due to Halcyon’s efficient beam utilization and fast gantry rotation design[22]. In contrast, TrueBeam showed a stronger linear correlation between PCI and MUs (R^2^ = 0.54), indicating that TrueBeam is more dependent on MUs when handling high-complexity plans, likely due to its single-layer HD-MLC design and dose optimization mechanism. These findings are valuable references for clinical selection of radiotherapy devices and the optimization of treatment plans.

This study makes significant academic contributions to the dosimetric performance comparison of radiotherapy devices. First, it systematically quantifies the performance differences between Halcyon and TrueBeam in terms of PTV dose distribution, OAR protection, and plan complexity, revealing Halcyon’s notable advantages in low-dose protection for the lungs, heart, and NT, and in target conformity, providing dosimetric evidence for equipment selection. Second, through correlation analysis, this study identifies the relationship between PTV volume changes and low-dose exposure to OARs, highlighting that as PTV volume increases, Halcyon provides more significant low-dose protection for patients, offering new insights for optimizing treatment plans for locally advanced cancer. Furthermore, the study reveals the differences in the correlation between plan complexity and MUs, suggesting that Halcyon demonstrates greater stability in handling complex plans. These findings provide new empirical evidence and theoretical support for optimizing radiotherapy equipment selection, improving treatment plan auto-optimization algorithms, and developing personalized treatment approaches.

Despite the valuable results, this study has limitations. First, the study only included IMRT techniques and did not compare other radiotherapy technologies such as volume-modulated arc therapy (VMAT), which may show significant differences in treatment efficiency, dose distribution, and MUs[23]. Second, this study analyzed radical treatment plans for NSCLC patients, without considering the applicability to other anatomical sites. Although this single-cancer study design helps control variables, it limits the generalizability of the findings to other cancers. Additionally, as a retrospective, single-center study, case selection may have introduced bias, which may not fully reflect the actual application in multi-center settings, limiting the external validity of the results. Finally, the potential relationship between plan complexity and machine monitor units has not been fully elucidated. Future studies could use more machine learning approaches to analyze the relationship between plan complexity, MUs, and treatment outcomes, providing smarter tools for radiotherapy optimization. Therefore, future research should expand to multi-center, multi-tumor site validations, introduce comparisons with VMAT, and explore the potential mechanisms behind plan complexity and MUs using data-driven approaches, thus providing more comprehensive evidence to support the clinical application of radiotherapy devices.

## Conclusion

This study systematically compared Halcyon and TrueBeam in terms of PTV dose distribution, OAR protection, and plan complexity. The results show that Halcyon has significant advantages in target conformity and low-dose protection (e.g., V_5_, V_10_ for the lungs and heart). This study also highlights differences in the dependence on plan complexity and machine monitor units, indicating that Halcyon shows greater stability and efficiency in handling complex plans. These findings provide essential references for clinical selection of radiotherapy equipment, emphasizing Halcyon’s potential value in reducing low-to mid-dose radiation exposure volumes for locally advanced lung cancer patients, and offer important insights for optimizing radiotherapy plans and equipment selection based on the individualized characteristics of lung cancer patients.

## Data Availability

All relevant data are within the manuscript and its Supporting Information files.

## Author Contributions

Shuangyan Yang: study design, statistical analysis, and manuscript writing; Yaping Xu: study design and manuscript revision.

